# Association between temporal muscle thickness and clinical outcomes in patients with newly diagnosed glioblastoma

**DOI:** 10.1101/2020.07.02.20145342

**Authors:** Geon An, Stephen Ahn, Jae-Sung Park, Sin-Soo Jeun, Yong-Kil Hong

**Affiliations:** Department of Neurosurgery, Seoul St. Mary’s Hospital, College of Medicine, The Catholic University of Korea, Seoul, South Korea

**Keywords:** Sarcopenia, Temporal muscle thickness, Glioblastoma, Cancer, Prognosis

## Abstract

**Purpose:** Temporal muscle thickness (TMT) has been suggested as a novel biomarker that can represent sarcopenia in head and neck malignancies. This study investigated the association of TMT with clinical outcomes in patients with newly diagnosed glioblastoma (GBM).

**Materials and Methods:** Using electronic medical records, all GBM patients between 2008 and 2018 at Seoul St. Mary’s Hospital were reviewed. Total 177 patients met our eligibility criteria.

**Results:** The thinner group who had TMT less than the median showed shorter overall survival (OS) and progression-free survival (PFS) than the thicker group who had TMT more than median (OS; 11.0 versus 18.0 months, *p* < 0.001, and PFS; 6.0 versus 11.0 months, *p* < 0.001). In the multivariate analysis, the thinner group had negative associations with OS and PFS (OS; HR 2.63 (1.34-2.63), *p* < 0.001, and PFS; HR 2.21 (1.34-2.50), *p* = 0.002). We also performed propensity score matching between the thinner and thicker groups to minimize the potential bias. The thinner group showed shorter OS and PFS (OS; 13.5 versus 19.0 months, *p* = 0.006, and PFS; 6.5 versus 9.0 months, *p* = 0.028) and had negative associations with OS and PFS than the thicker group (OS; HR 1.90 (1.19-3.03), *p* = 0.008, and PFS; HR 1.70 (1.07-2.70), *p* = 0.026) in matched patients.

**Conclusion:** Our findings suggest that TMT can be a useful prognostic biomarker for clinical outcomes in GBM patients. Further preclinical and clinical studies could help elucidate this association of sarcopenia with clinical outcomes in GBM patients.

## Introduction

Glioblastoma (GBM) is the most common and most aggressive primary brain tumor in adults[1]. The standard treatment for GBM includes maximal and safe surgical resection and concomitant chemoradiation (CCRT) followed by 6 cycles of adjuvant temozolomide (TMZ) chemotherapy[2]. Despite these aggressive multidisciplinary treatments, the median survival is less than 15 months, with a 2-year survival rate < 30% and a 5-year survival rate < 10%[3].

Sarcopenia, defined as loss of skeletal muscle mass, is known to be a poor prognostic factor in various solid cancers[4, 5]. Skeletal muscle mass is usually measured by the skeletal muscle cross-sectional area at the level of the adjacent lumbar vertebrae on computed tomography (CT) scan[6, 7]. However, measuring skeletal muscle mass is not feasible in most clinics of neuro-oncology because abdominal CT scans are not routinely performed[8]. For this reason, evidence to support sarcopenia as related to clinical outcomes of brain tumor patients has been relatively limited compared with those of other cancers.

Recently, several studies suggested that temporal muscle thickness (TMT), which is easily obtained on routine brain MRI, was highly related to lumbar skeletal muscle mass[9, 10] and could be a novel biomarker that represents sarcopenia in head and neck malignancies[11, 12], including brain metastasis[13, 14]. To date, only one recently published study has supported this association in GBM[15].

In this study, we tried to determine if TMT could be an independent prognostic factor for OS and PFS in patients with newly diagnosed GBM. We retrospectively analyzed the prognostic relevance of TMT with previously well-known factors including age, extent of resection, and performance status of GBM patients.

## Materials and Methods

### Patient population

This retrospective study was approved by the Institutional Review Board of Seoul St. Mary’s Hospital. After approval, the electronic medical records of newly diagnosed GBM patients treated at Seoul St. Mary’s Hospital between 2008 and 2018 were examined. The eligibility criteria were: 1) newly diagnosed primary GBM, 2) pathologically confirmed by craniotomy or stereotactic biopsy, 3) accessible baseline thin slice magnetic resonance imaging (MRI), and 4) accessible survival status and/or death date. The exclusive criteria were: 1) prior history of brain surgery or radiation therapy due to any medical illness or trauma, 2) proven molecular features with IDH mutation, and/or 1p19q co-deletion. A summary of patient enrollment is illustrated in Figure 1.

**Figure 1.**
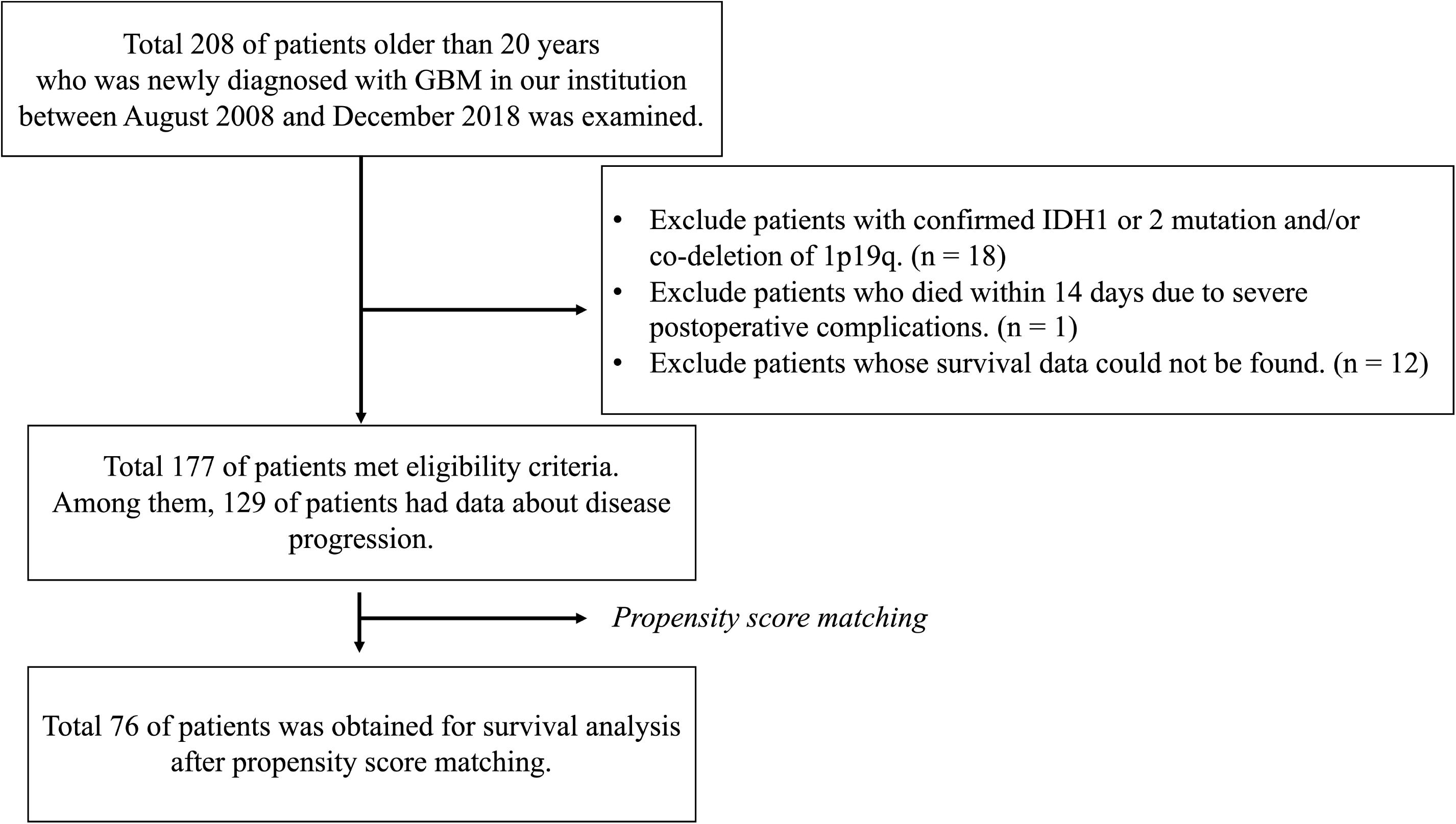
Study design.

### Temporal muscle thickness (TMT)

TMT was calculated on axial thin slice (1mm cut) contrast-enhanced T1-weighted MR images, which was routinely performed on the surgery day for navigation. The plane was oriented parallel to the anterior commissure-posterior commissure line. The measurements were performed perpendicular to the long axis of the temporal muscle using the orbital roof and the Sylvian fissure as anatomical landmarks, according to previously reported methods[13-15]. The TMT of the left and right side was measured separately in each patient, and the TMT of each side was summed and divided by two, which resulted in a mean TMT. Examples of TMT measurements on brain MRI are shown in Figure 2.

**Figure 2.**
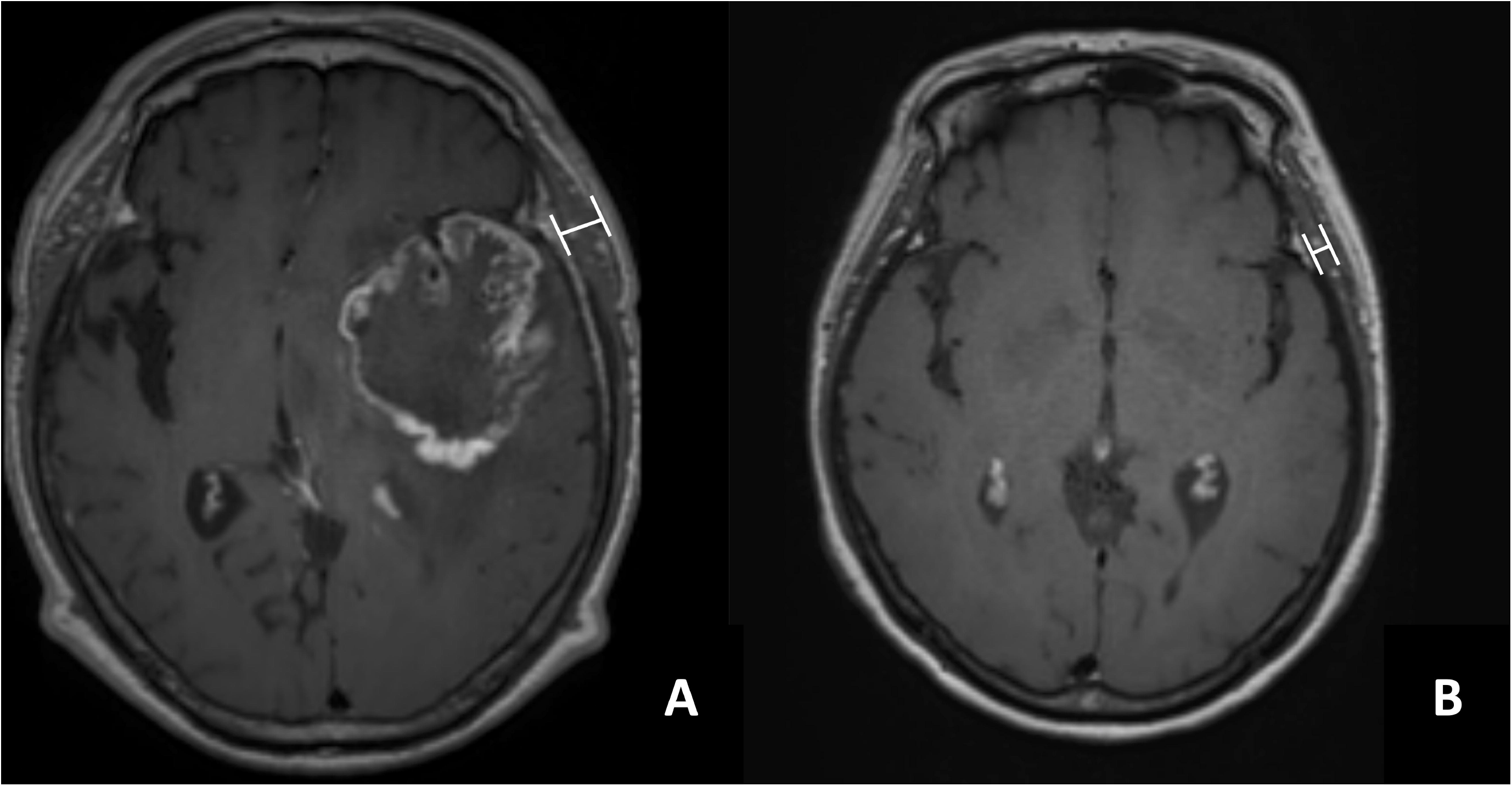
TMT measurement on brain MRI. (a) A 32-year-old male patient with an overall survival of 15 months (Mean TMT = 12.3 mm) and (b) a 73-year-old female patient with an overall survival of 2 months (mean TMT = 5.5 mm).

### Clinical variables

Clinical variables of sex, age, extent of surgical resection (EOR), pathological diagnosis, molecular features, dose and fraction of radiation, type of chemotherapy and number of cycles administered, radiological findings, and status of survival and/or death date were collected. The EOR was measured by comparing radiologic findings on MRI at baseline and within 48 hours after surgery. Resection of 90% of the tumor volume was defined as GTR, and resection of <90% was defined as non-GTR, according to previous studies[16, 17]. After surgical resection, we tried to start concomitant chemoradiotherapy (CCRT) within 28 days. The radiation dose was 5940 cGy for 33 fractions or 6000 cGy for 30 fractions. TMZ dose was 75 mg/m^2^ during CCRT and 150-200 mg/m^2^ during 6 cycles of adjuvant therapy, regardless of age. Isocitrate dehydrogenase 1 (IDH) mutation was evaluated by immunohistochemistry or directing sequencing. If necessary, IDH 2 mutation was evaluated by directing sequencing. The presence of 1p19q co-deletion was examined using fluorescence in situ hybridization (FISH). The O^6^-methylguanine-DNA methyltransferase (MGMT) gene methylation status was evaluated by polymerase chain reaction. Status of survival and/or death dates were obtained from the Korea Central Cancer Registry database. The OS was defined as days from initial surgery to death, and PFS was defined as days from initial surgery to progression confirmed by MRI, according to response assessment in neuro-oncology criteria (RANO). Patients who were confirmed to be alive on December 31, 2019, were censored. The average duration of follow-up was 14.0 months (range, 1-123 months).

### Statistical analysis

The median value of TMT in each male and female patient was determined as the TMT cut-off point based on previous studies. All clinical variables were considered with descriptive statistics. The differences between groups were compared using Fisher’s exact test or the chi-square test. The normality test was performed for continuous variables. Kaplan-Meier survival analysis and the log-rank test were used to estimate median OS and PFS. Univariate and multivariate analyses were conducted using a Cox proportional regression model. Hazard ratios (HRs) and 95% confidence intervals (CIs) were calculated. Multivariate analysis was performed on the variables with P values <0.2, and P values <0.05 were considered to indicate statistical significance.

Propensity score matching (PSM) was performed to decrease potential bias and balance the baseline variables between the two groups. Matching was performed using clinical variables that were likely associated with prognostic factors including sex, age, extent of surgical resection, and performance score[18]. Analysis was performed with 1 to 1 nearest neighbor matching with replacement and a caliper size of 0.1 standard deviation. An absolute standardized difference (SMD) less than <0.20 was considered a negligible imbalance, and SMD of all clinical variables was reduced to less than 0.2 after matching. All statistical analysis was estimated using R Statistical Software (Version 3.2.3).

## Results

### Patient Characteristics

A total of 177 patients who met the eligibility criteria was included. The overall median TMT of patients was 6.26 mm (range 2.41–12.48), that for male patients was 7.10 mm (range 3.55– 11.68), and that for female patients was 5.54 mm (range 2.41–12.48) (p < 0.001). We set the cut-off point as the median value of each male and female patient group. Eighty-eight patients who had TMT less than the median were assigned to the “thinner group,” while 89 patients who had TMT greater than the median was assigned to the “thicker group.” The baseline characteristics of these groups are described in Table 1. Patients with GTR or good performance were more prominent in the thicker group, while older patients were more prominent in the thinner group.

**Table 1.**
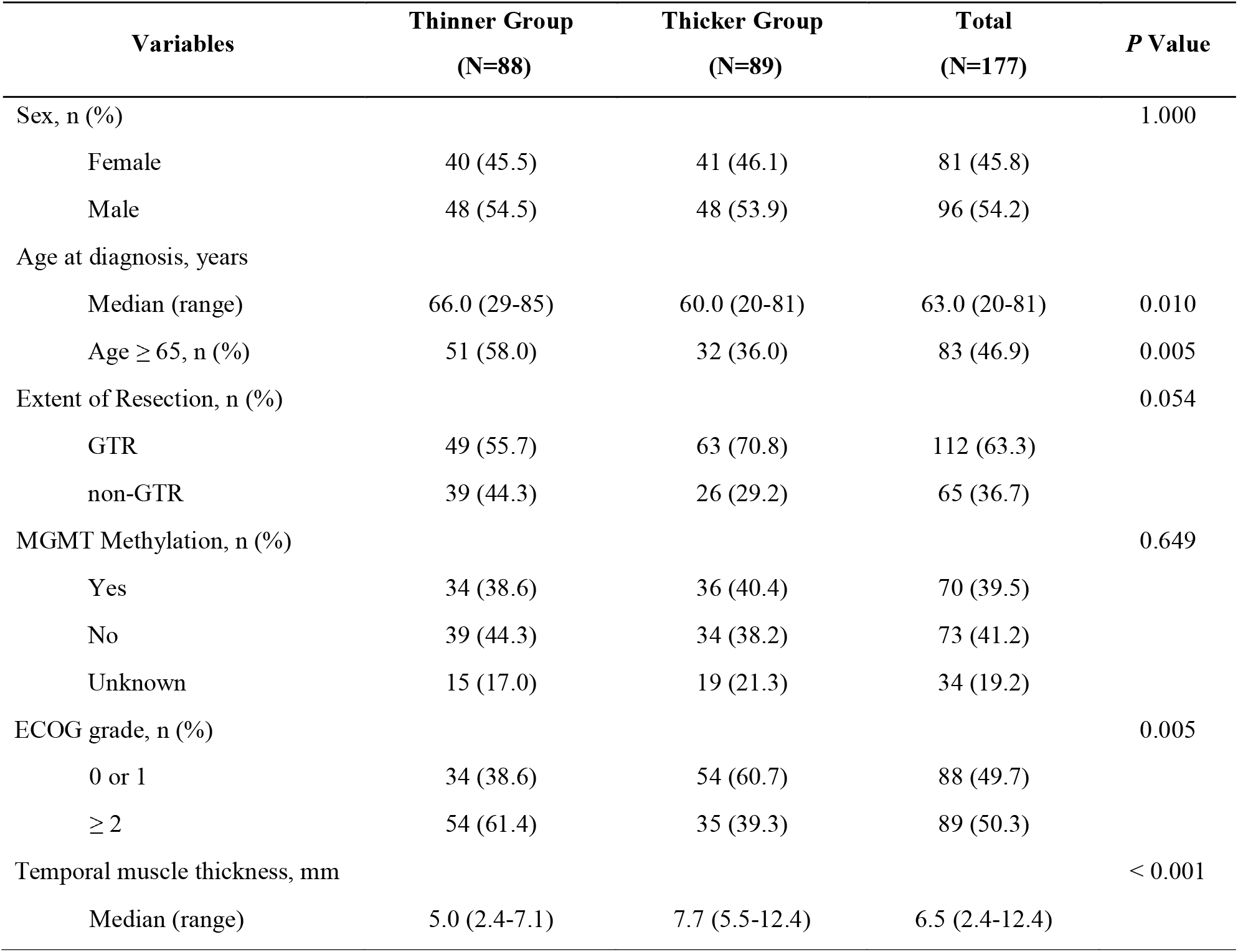
Patient characteristics.

### TMT and Clinical Outcomes

We used a Kaplan-Meier survival analysis and multivariate Cox regression analysis with previously known prognostic factors to evaluate whether TMT was associated with OS and PFS. The Kaplan–Meier survival curves of OS and PFS for the thicker and thinner groups are illustrated in Figures 3a and 3b. The median OS of the thicker group was longer than that of the thinner group (18.0 months versus 10.0 months, *p* < 0.001). The median PFS of the thicker group was also longer than that of the thinner group (11.0 months versus 6.0 months, *p* < 0.001).

**Figure 3.**
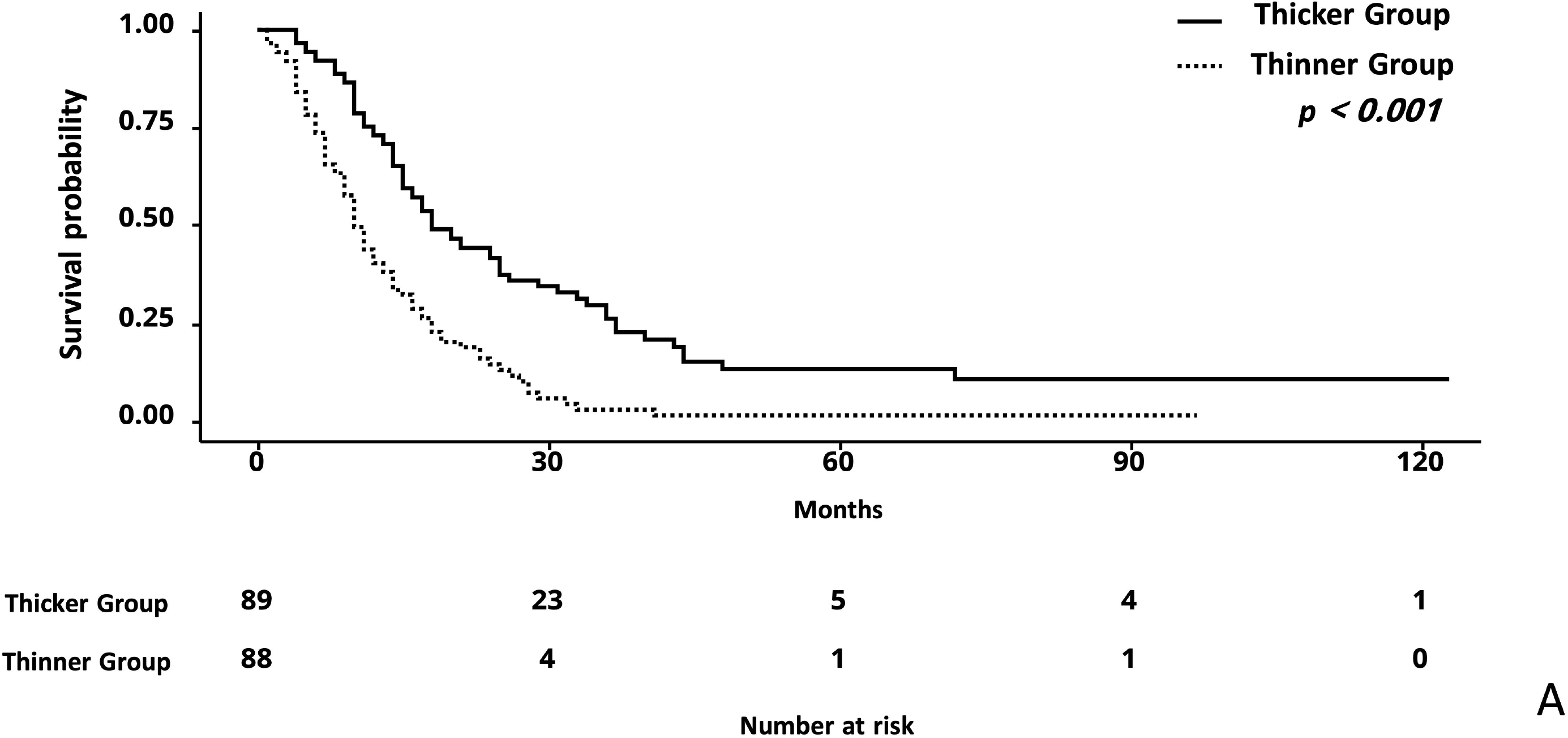
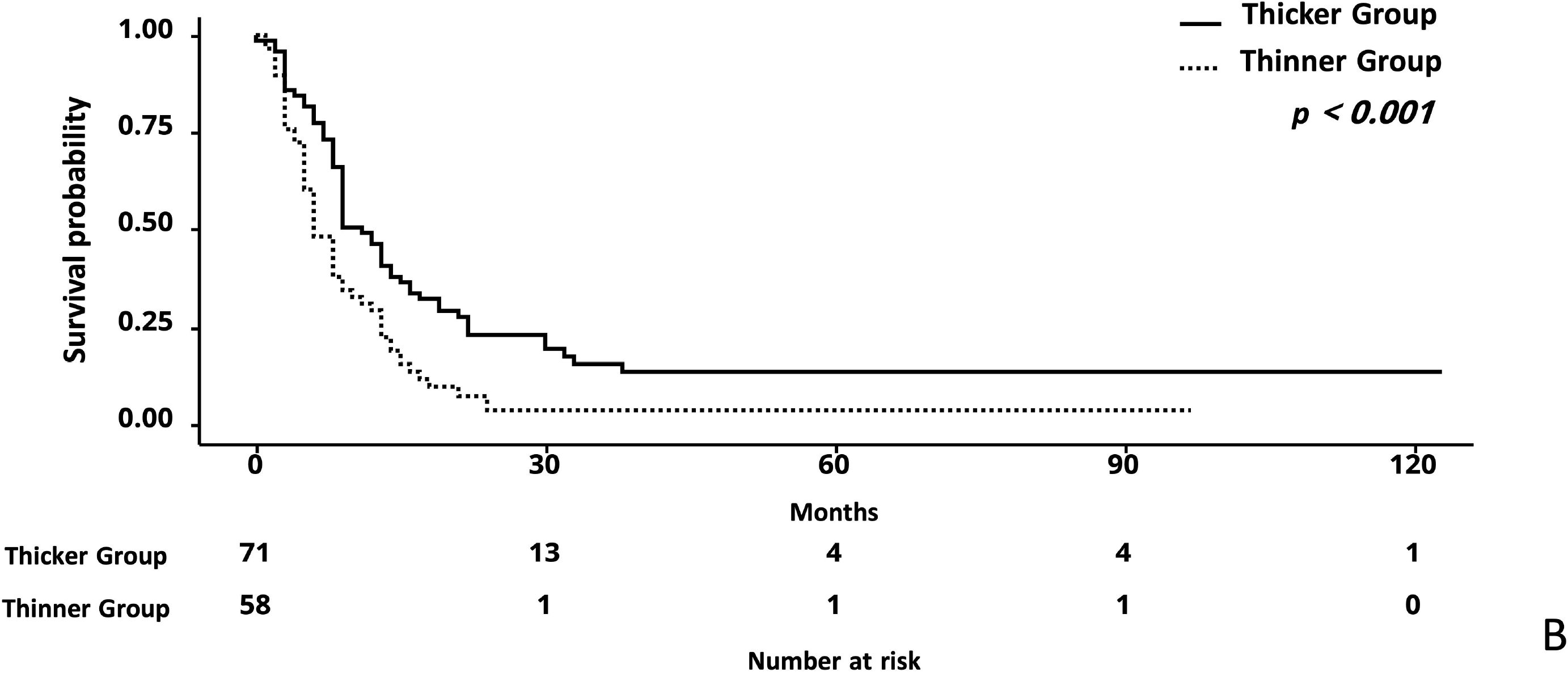
Kaplan-Meier survival curve for (a) overall survival and (b) progression free survival comparing the thinner group with the thicker group.

Univariate and multivariate Cox analyses for OS and PFS are described in Table 2 and Table 3. In multivariate analysis for OS, a thinner TMT (less than the median) was negatively associated with OS (HR 2.63 CI 1.34-2.63, *p* < 0.001). Non-GTR (HR 1.75 CI 1.22-2.50, *p* = 0.002) and an ECOG of 2 or 3 (HR 2.21 CI 1.51-3.24, *p* < 0.001) were negatively associated with OS, while sex and age ≥ 65 were not associated with OS in this study. Similar results were obtained in multivariate analysis for PFS. A thinner TMT (less than the median) was negatively associated with PFS (HR 1.74 CI 1.14-2.65, *p* = 0.009). Non-GTR (HR 1.75 CI 1.22-2.50, *p* = 0.002) and an ECOG of 2 or 3 (HR 2.41 CI 1.58-3.68, *p* <0.001) were negatively associated with shorter PFS, while sex and age ≥ 65 were not associated with PFS in this study.

**Table 2.**
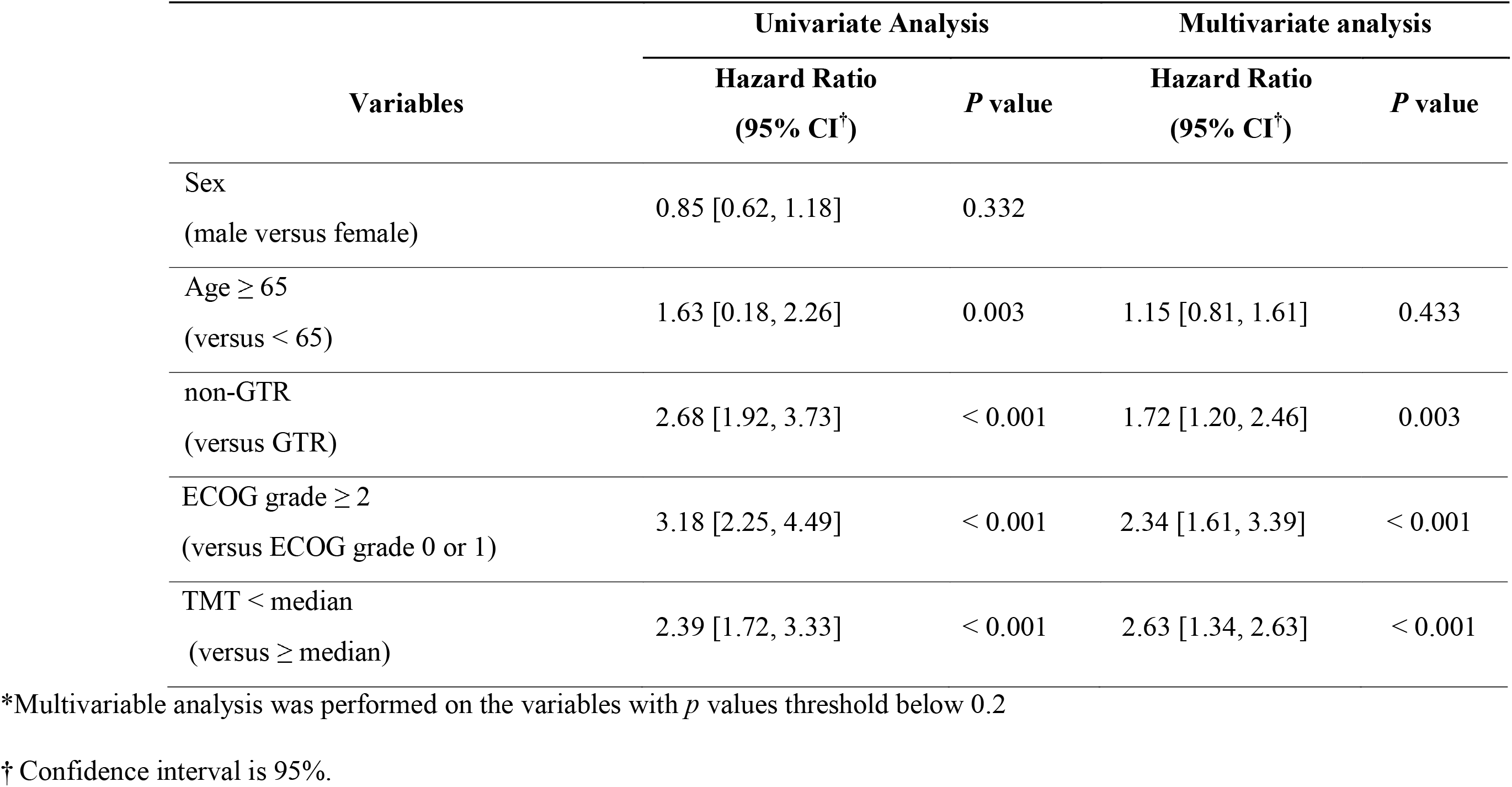
Univariate and multivariate Cox regression analysis for overall survival.

**Table 3.**
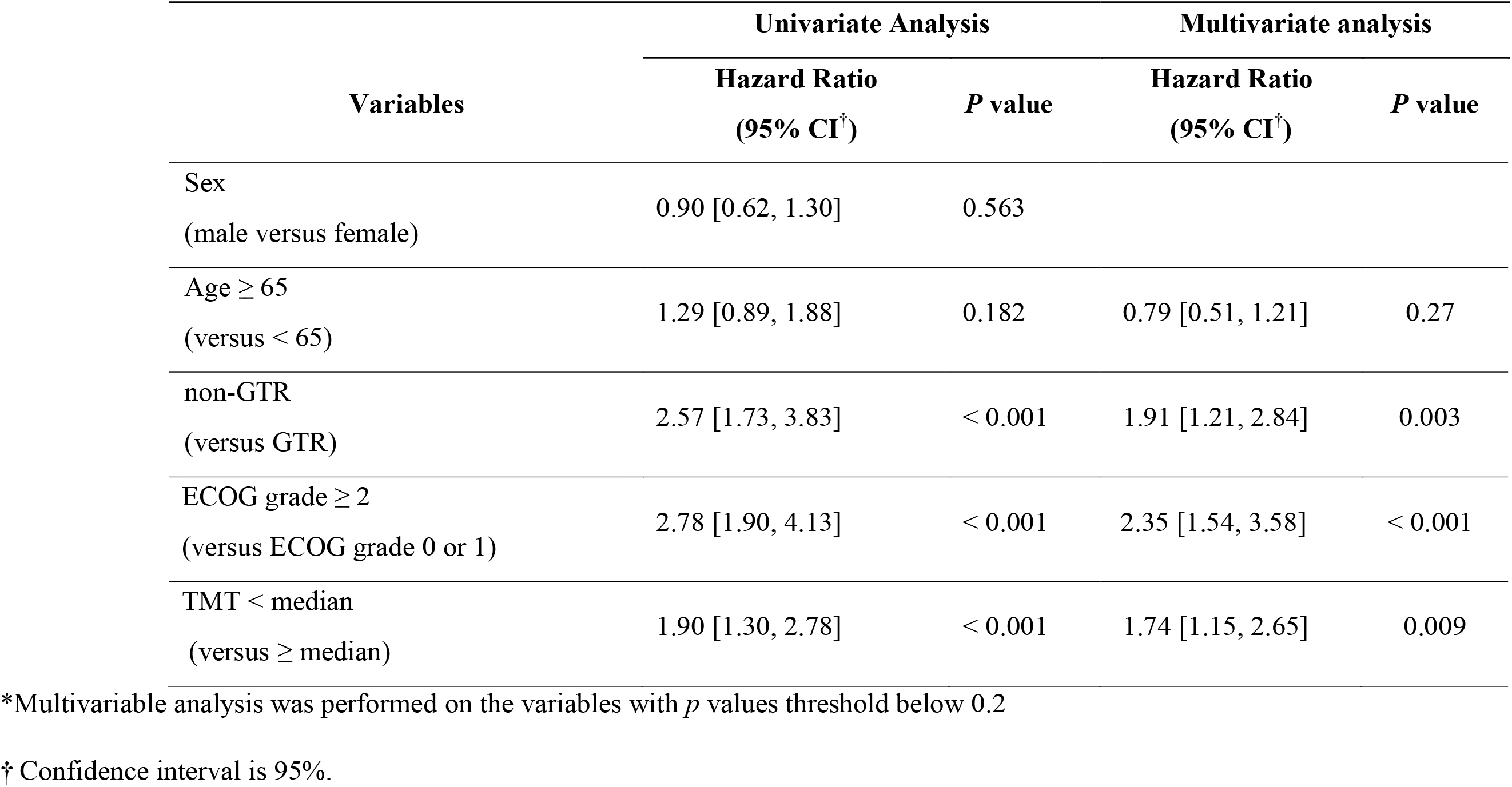
Univariate and multivariate Cox regression analysis for progression free survival.

### Propensity Score Matching Analysis

Because the baseline characteristics above, and described in Table 1, were not homogeneous between the two groups, we performed PSM between the thinner and thicker groups to minimize the potential bias. Age, sex, EOR, and ECOG were included as co-variables. After matching theses variables, the difference of variables between the matched patients in the thicker and thinner groups was not statistically significant, and the detailed information is described in Table 4. The median OS of the thinner and thicker groups was 13.5 and 19.0 months, respectively (*p* = 0.006). The median PFS of the thinner and thicker groups was 6.5 and 9.0 months, respectively (*p* = 0.028). The HR of the thinner TMT (less than the median) for OS was 1.90 (CI 1.19-3.03, *p* = 0.008). The HR of the thinner TMT (less than the median) for PFS was 1.70 (CI 1.07-2.70, *p* = 0.026). The Kaplan-Meier survival curves for OS and PFS are illustrated in Supplementary Figures 1a and 1b.

**Table 4.**
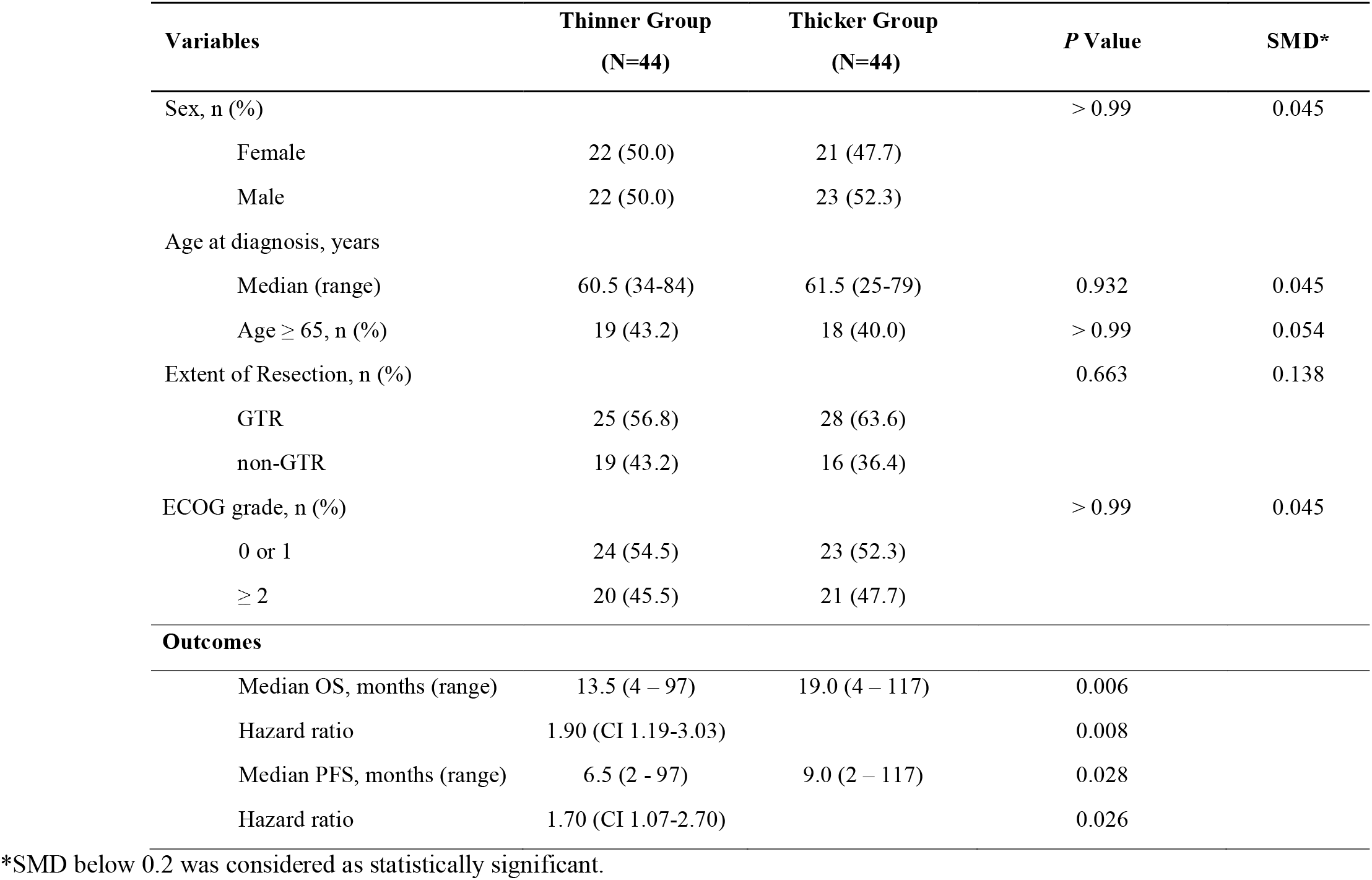
Propensity score matching between thicker and thinner groups and comparing of clinical outcomes between thicker and thinner groups.

## Discussion

Numerous studies have supported the hypothesis that body composition is associated with clinical outcomes in various cancers[7, 19-23]. Sarcopenia, defined as loss of skeletal muscle mass, has specifically been suggested as a significant and independent biomarker for clinical outcomes, postoperative complications, and toxicity induced by chemotherapy in various cancer patients[11, 24-26]. Skeletal muscle mass is usually measured by the total cross-sectional skeletal muscle area at the level of the third lumbar vertebrae, using a CT scan[6, 7]. When evaluating radiologic findings of cancers, such as esophageal cancers, colorectal cancers, and pancreatic cancers, images that are needed to calculate skeletal muscle mass could be routinely acquired. However, in head and neck cancers, including brain tumors, images that are needed to calculate skeletal muscle are not feasible in clinics. Therefore, the evidence to support whether sarcopenia was associated with brain tumors has been limited and the results have not been conclusive.

Several recent studies have suggested that TMT could be easily acquired and could be an alternative biomarker for sarcopenia[9, 10, 13-15]. Ranganathan et al. suggested that TMT was correlated with the psoas muscle and was associated with clinical outcomes in trauma patients[10]. Leitner et al. found that TMT was correlated with skeletal muscle mass obtained at the lumbar level and had independent prognostic relevance in lung cancer and in melanoma patients with brain metastasis[9]. Further et al. also showed that TMT was an independent prognostic marker in brain metastasis patients[13, 14]. A recent study published by Furhter et al. found that TMT was also an independent prognostic marker in patients with progressive GBM[15].

The pathophysiological mechanism of the association between sarcopenia and poorer outcomes in cancer patients has remained relatively uncertain. Recent findings suggest five possible mechanisms: the role of skeletal muscle mass to modulate inflammation through the immune system via cytokines and myokines, the influence on insulin-dependent glucose control, mitochondrial function, synthetic and degradative protein pathways, and pharmacokinetics of anticancer drugs[27]. In this context, various therapeutic approaches to restore sarcopenia have been attempted[28]. Physical exercise may help to prevent skeletal muscle decline by stimulating myokine production, which may prevent negative effects on clinical outcomes[29]. Additionally, supplements including omega-3 fatty acid, melanocortin-4 receptor antagonists, myostatin inhibition, and IL-6 antagonism are being currently investigated as novel methods to restore sarcopenia[30-33].

In this framework, we tried to validate the hypothesis that sarcopenia can affect clinical outcomes of newly diagnosed GBM patients, like other solid cancers, and that TMT could be an independent and objective biomarker for sarcopenia as an alternative for skeletal muscle mass. We set different cut-off points of TMT for each male and female patient, according to previous studies. In this study, the medial TMT was different between male and female patient groups (7.10 mm vs 5.54 mm, p < 0.001). The results showed that thinner TMT was an independent prognostic factor for OS (HR 2.63 CI 1.34-2.63, *p* < 0.001) and PFS (HR 1.74 CI 1.15-2.65, *p* = 0.009) in patients with newly diagnosed GBM. Because baseline characteristics including age, EOR, and performance scores were different between the two groups, we performed PSM to minimize potential bias. After matching co-variables of sex, age, EOR, and performance scores, which could affect clinical outcomes, we showed that thinner TMT had negative associations with OS (HR 1.90 CI 1.19-3.03, *p* = 0.008) and PFS (HR 1.70 CI 1.07-2.70, *p* = 0.026) than thicker TMT in this matched population.

Our findings are consistent with previous studies which evaluated if TMT could be related with sarcopenia and if TMT could be related with clinical outcomes in brain malignancies[9, 14, 15]. Taken together, we suggest that TMT as a surrogate marker for sarcopenia could be an independent prognostic factor for clinical outcomes in patients with newly diagnosed GBM. Also, when physicians consider patient’s physical condition for determining treatment modalities and their intensity, TMT also may be suggested as alternative biomarkers that represent patient’s physical status in addition to generally well-considered factors such as age and performance status.

Our findings should be considered in light of several limitations. First, selection bias is possible due to the retrospective study approach. Second, molecular parameters, such as IDH1, 1p19q co-deletion, and MGMT gene methylation status, were not fully defined. Third, the baseline characteristics, especially EOR, were significantly different between the two groups, although we verified that TMT was an independent significant prognostic factor for both OS and PFS in multivariate Cox regression analysis; we also matched the baseline characteristics and showed that TMT was a significant prognostic factor for OS and PFS in these matched groups. Findings that sarcopenia is common in elderly patients may explain the higher incidence of younger patients, receiving of GTR, or good performance status in the thicker group[34]. Because younger patients usually show better performance and are more likely to undergo more aggressive surgery that could help them achieve maximal resection compared with older patients[35]. Due to several limitations of our study, further prospective studies and preclinical studies are needed to validate our findings and analyze the associations between TMT and EOR.

In summary, our findings showed that TMT could be an independent prognostic marker for OS and PFS in patients with newly diagnosed GBM. The results also supported that sarcopenia could be related to GBM in patients.

## Data Availability

All data supporting the fnidings presented in this manuscript are available upon reasonable request directly to the corresponding author

## Figure legends

Supplementary Figure 1. Kaplan-Meier survival curve for (a) overall survival and (b) progression free survival comparing the thinner group with the thicker group in a matched population using propensity score matching.

